# Metabolic alterations in human post-mortem frontal cortex and cerebrospinal fluid associated with high levels of nicotine metabolite cotinine

**DOI:** 10.1101/2025.01.24.25321062

**Authors:** Wadzanai Masvosva, Marko Lehtonen, Mika Martiskainen, Jari Tiihonen, Pekka J. Karhunen, Kati Hanhineva, Jaana Rysä, Eloise Kok, Olli Kärkkäinen

## Abstract

Cigarette smoking is the single most significant cause of preventable death in the world. Tobacco smoking causes exposure to thousands of chemicals and disrupts biological pathways. It impacts several organs, including the brain, where its effects are mediated by nicotinic acetylcholine receptors. Women seem to be more susceptible to the negative health effects of smoking. In this study we focused on the changes in the metabolic profile of human postmortem frontal cortex and cerebrospinal fluid samples associated with high levels of the nicotine metabolite cotinine. We used non-targeted metabolomics to analyze post-mortem frontal cortex and cerebrospinal fluid (CSF) samples from the Tampere Sudden Death Study cohort. We identified 137 cases (24 females) with high cotinine levels, indicating nicotine exposure. For controls, we identified 82 subjects (20 females) with no cotinine in the frontal cortex or CSF samples and no known history of smoking based on medical records and autopsy reports. Cases had significantly higher levels of 1-methylhistamine (Cohen’s d=0.66, p<0.0001) and N-acetylputrescine (d=0.84, p<0.0001), and lower levels of aspartic acid (d=-0.53, p<0.001), 3-methylhistidine (d=-0.58, p=0.0004), and taurine (d=-0.47, p=0.0002) in the frontal cortex compared to controls. Compared to the frontal cortex, differences between cases and controls were smaller in the CSF samples. Most of the observed differences were similar in both sexes, with a few exceptions like low ergothioneine levels, observed especially in female cases. In conclusion, smoking or nicotine exposure is associated with alterations in metabolites linked to increased oxidative stress and neuroinflammation, as well as reduced neurotransmitter levels in the frontal cortex.

## Introduction

Cigarette smoking is the single most significant cause of preventable death in the world^1^. Tobacco smoking causes exposure to thousands of chemicals and can disrupt several biological pathways, leading to negative health effects. A key component in tobacco is nicotine, which is considered to be the main cause of addiction to tobacco smoking and is currently also used in e-cigarettes, nicotine replacement products, and other tobacco-free nicotine products^2^. For example, nicotine is an important factor in the initiation and progression of smoking-induced atherosclerosis^3^. Nicotine also dysregulates apoptosis signaling pathways, which in turn initiates lung cell apoptosis and contributes to lung diseases^4^. In human metabolism, nicotine is converted to cotinine that can be used to assess recent nicotine exposure^5^.

Besides the heart and lungs, the brain is a target organ for nicotine, where its effects are mediated by nicotinic acetylcholine receptors^6^. Repeated long-term exposure to nicotine alters the function of the brain. For example, it increases the levels of nicotinic acetylcholine receptors, which in turn affects other neurotransmitter systems, including glutamatergic and GABAergic systems^7^. Neuroimaging and neuropathological investigations show a wide range of effects of nicotine exposure on the brain. Nicotine exposure has also been associated with damage to the endothelial cells, resulting in an increased risk of cerebrovascular disease^8^. Smoking has been connected to structural decreases in brain regions such as the right cerebellum, multiple prefrontal cortex regions and the thalamus. This decrease in gray matter seems to have structural-behavioral implications, such as visual processing problems in the prefrontal cortex^9^.

Moreover, some of the effects of nicotine on the brain seem to differ between sexes^10^. For example, when exposed to tobacco smoking, males show reduced left caudate volume with altered functional connectivity, whereas females show a decreased right amygdala volume and altered impulsivity correlations^11^. However, possible sex differences in the metabolic pathways of human brain are not known.

Non-targeted metabolomics is an excellent tool for measuring the effects of complex chemical exposures, such as tobacco smoking. Non-targeted metabolomics measures hundreds of endogenous, microbiota associated, and exogenous small molecules in a sample and can therefore provide wide understanding of metabolic adaptations related to different exposures^12^. Smoking is associated with alterations in the circulating metabolite profile^13–17^. Some of these changes could be important for brain function and may be associated with nicotine dependence. For example, both tobacco smoking and use of heated tobacco products have been linked with high blood levels of glutamate, the main excitatory neurotransmitter^14,16^. On the other hand, reduced blood levels of glutamate seem to be an acute effect of cigarette smoking in current smokers^15^. Furthermore, levels of another neurotransmitter, serotonin, has been reported to be increased in the blood in current smokers^13^. However, circulating levels might not reflect levels in the brain^18^. Understanding how tobacco smoking or nicotine exposure alters the brain metabolome could be important for understanding brain related pathology associated with tobacco, including nicotine dependence. However, no metabolomics studies have been performed to investigate the effects of tobacco smoking or nicotine exposure on human brain tissue.

Here, our aim was to investigate changes in the human brain and cerebrospinal fluid (CSF) metabolite profiles associated with tobacco smoking or nicotine exposure. We conducted a non-targeted metabolomics analysis using post-mortem frontal cortex and CSF samples to gain novel insights into tobacco smoking or nicotine exposure-related changes in the metabolic processes of the human brain. We also investigated possible sex-dependent associations in the post-mortem samples.

## Methods and Materials

### Subjects

We used post-mortem frontal cortex and CSF samples from the Tampere Sudden Death Study (TSDS) cohort^19^. The samples were collected from forensic autopsies on people who suffered out-of-hospital deaths in the area of the Pirkanmaa Hospital District (Finland) during 2010-2015. Frontal cortex (Broadman area 9) and CSF samples, a portion extracted via syringe, were collected from a total of 700 subjects and stored at -80 °C until use.

The inclusion criteria for the current study cases were high cotinine (metabolite of nicotine) levels in the CSF and frontal cortex samples. Smoking status, based on medical and autopsy records, was missing from 64 cases (47%). Of those with a known smoking history, most were tobacco smokers (69, 95%) and the rest smoked pipes or cigars. No cotinine in the samples and no known history of smoking were inclusion criteria for the control group. We excluded seven subjects who did not show smoking in medical records, but did have high cotinine levels in frontal cortex or CSF samples. Furthermore, we excluded one subject who was classified as a smoker based on medical records, but did not have any cotinine in the frontal cortex or CSF samples. In the end, we had 137 subjects in the smoking group and 82 in the control group (Table 1).

**Table 1:**
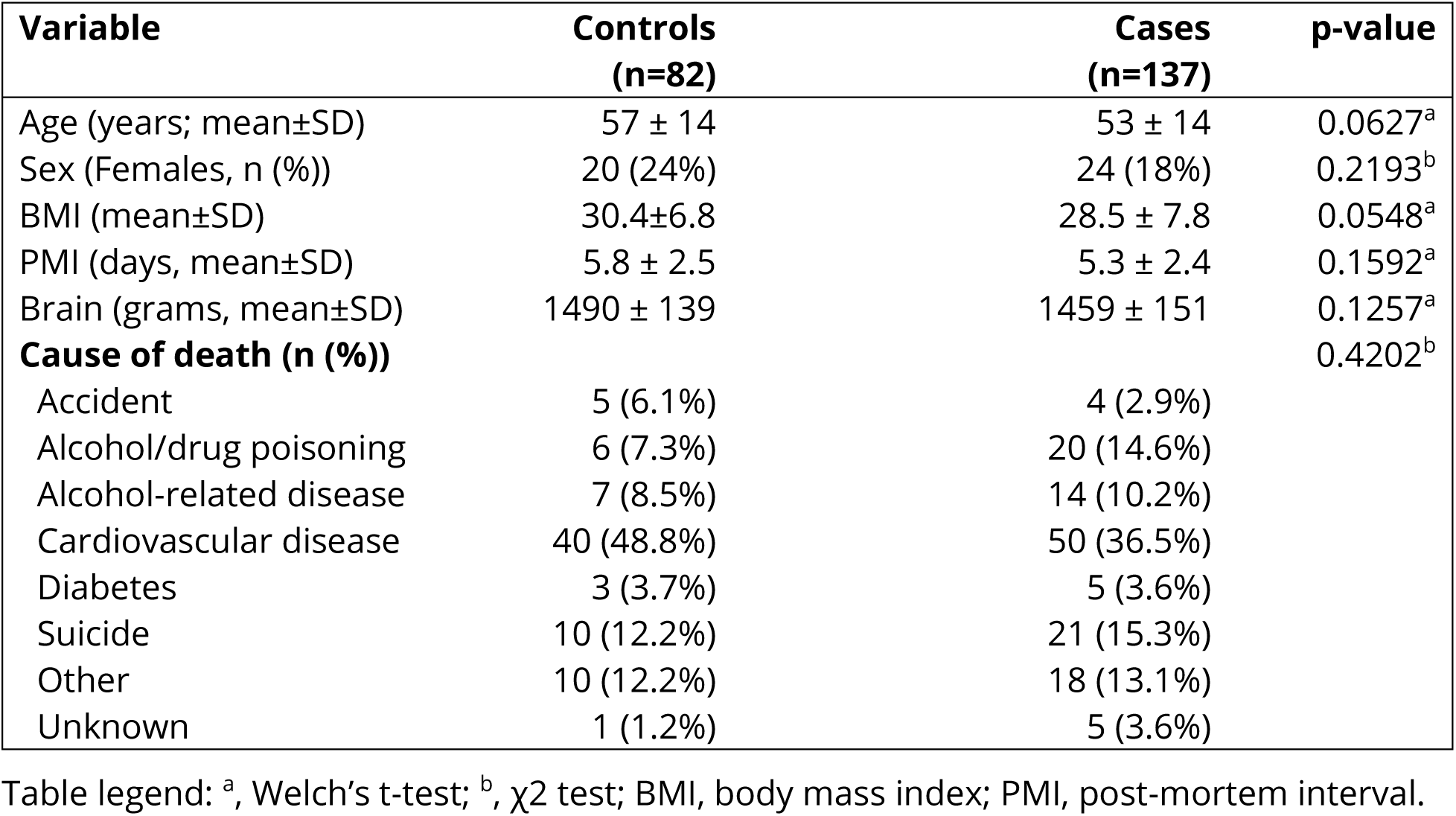
Background characteristics.

| Variable | Controls<br>(n=82) | Cases<br>(n=137) | p-value |
| --- | --- | --- | --- |
| Age (years; mean±SD) | 57 ± 14 | 53 ± 14 | 0.0627<sup>a</sup> |
| Sex (Females, n (%)) | 20 (24%) | 24 (18%) | 0.2193<sup>b</sup> |
| BMI (mean±SD) | 30.4±6.8 | 28.5 ± 7.8 | 0.0548<sup>a</sup> |
| PMI (days, mean±SD) | 5.8 ± 2.5 | 5.3 ± 2.4 | 0.1592<sup>a</sup> |
| Brain (grams, mean±SD) | 1490 ± 139 | 1459 ± 151 | 0.1257<sup>a</sup> |
| **Cause of death (n (%))** |  |  | 0.4202<sup>b</sup> |
| Accident | 5 (6.1%) | 4 (2.9%) |  |
| Alcohol/drug poisoning | 6 (7.3%) | 20 (14.6%) |  |
| Alcohol-related disease | 7 (8.5%) | 14 (10.2%) |  |
| Cardiovascular disease | 40 (48.8%) | 50 (36.5%) |  |
| Diabetes | 3 (3.7%) | 5 (3.6%) |  |
| Suicide | 10 (12.2%) | 21 (15.3%) |  |
| Other | 10 (12.2%) | 18 (13.1%) |  |
| Unknown | 1 (1.2%) | 5 (3.6%) |  |

Table legend: <sup>a</sup>, Welch's t-test; <sup>b</sup>, $\chi^2$ test; BMI, body mass index; PMI, post-mortem interval.

### Non-targeted metabolomics analysis

The LC-MS analysis of the frontal cortex and CSF samples has been described in detail by Kärkkäinen et al (2021)^19^. In brief, tissue samples were weighed with an added 80% methanol (v/vH2O, LC-MS Ultra CHROMASOLV®, Fluka). 1000 μL solvent was added for each 100 mg of frontal cortex tissue. For the CSF samples, 400 μl of acetonitrile (VWR International, LC-MS grade) was added to 100 µL of CSF and mixed by pipette. Quality control samples were made by taking 5 μL solution of each sample. The samples were analyzed using a UHPLC-qTOF-MS system (Agilent Technologies, Waldbronn, Karlsruhe, Germany). It consisted of a 1290 LC system, a jet stream electrospray ionization (ESI) source and 6540 UHD accurate mass qTOF spectrometer. Samples were analyzed using two chromatographic techniques, reversed-phase (RP, Zorbax Eclipse XDB-C18, particle size 1.8 μm, dimensions of 2.1 × 100 mm, Agilent Technologies, USA) and hydrophilic interaction (HILIC, Acquity UPLC BEH Amide 1.7 μm,2.1 × 100 mm, Waters, Ireland), with both positive and negative ionization. Quality control samples were injected at the beginning of the analysis and at every 12^th^ sample. The analysis order of the samples was randomized. Three different collision-induced dissociation voltages were used (10, 20, and 40 eV) for the data-dependent tandem mass spectrometry (MSMS) analysis.

Metabolomics data-analysis was performed using MS-DIAL ver. 3.40^20^ and Notame R-package^21^. The peak picking and peak alignment parameters were: 1) mass range from 40 to 1000 (HILIC) or 1600 (RP) Da, MS tolerance 0.005 Da, MS2 tolerance 0.01 Da, minimum peak width 10 scans, minimum peak height 10,000 (selected based on background noise level). Peaks needed to be detected in at least 70% of samples from one study group to be included in the final data matrix. Drift correction of the metabolomics data was done using results from the QC sample, according to a previously published protocol^21^. In brief, the molecular features were corrected for the drift pattern caused by the LC-MS procedures using regularized cubic spline regression, fit separately for each feature on the QC samples. The smoothing parameter was chosen from an interval between 0.5 and 1.5 using leave-one-out cross-validation to prevent overfitting. After the drift correction, feature quality was assessed, and low-quality features were flagged. Features were kept if their RSD* was below 20% and their D-ratio below 40%. In addition, features with classic RSD, RSD*, and basic D-ratio all below 10% were kept. This additional condition prevents the flagging of features with very low values in all but a few samples. Missing values were imputed using random forest imputation. QC samples were removed prior to imputation to prevent them from affecting the imputation.

Metabolite identification was focused on statistically significantly altered (see “statistical analysis”) molecular features. Metabolite identification was based on exact mass, isotopic pattern, MS/MS fragmentation and retention time. Identifications were ranked according to community guidelines^22^. Metabolites at level 1 were matched to an in-house library built with chemical standards using the same experimental conditions. Level 2 includes metabolites matching the exact mass and MSMS spectra from public libraries (METLIN, LipidMaps, and Human Metabolome Database were used) or, in the case of lipids, the built-in MS-DIAL library version 3.40. In level 3, only the chemical group of the compound (but not the exact compound) is identifiable. Level 4 indicates unidentified compounds.

### Statistical analysis

We used Welch’s t-test (continuous variables) and χ² test (binomial variables) to evaluate differences between the study groups in background characteristics. We used Welch’s t-test and Cohen’s d-effect sizes to evaluate differences between the study groups in the metabolomics data. To estimate the possible role of sex, we performed additional analysis separately for both sexes. To account for multiple testing, we adjusted the α level by the number of principal components needed to explain 95% of the variation in the data. Here, 133 principal components were needed, and therefore the α level was adjusted to 0.0004. P-values between 0.05 and 0.0004 were considered trends. For multivariate analysis, we used partial least squares discriminant (PLS-DA) analysis using SIMCA (v 17.0, Sartorius Stedim Data Analytics Solutions) to calculate variable importance to projection (VIP) values. Identified metabolites with p-value < 0.05 were correlated with PMI using Pearson’s method.

## Results

The background characteristics (age, sex distribution, body mass index [BMI], post-mortem interval [PMI], brain weight and causes of death) were similar between the study groups. The cases in comparison to the controls, tended to be younger and slightly less overweight, however, these differences did not reach statistical significance (Table 1).

In the metabolomics analysis, both frontal cortex and CSF samples showed differences between the study groups in metabolite profiles (Figures 1-3, Supplementary Table S1). As per the selection criteria, the nicotine metabolite cotinine was significantly higher in both frontal cortex and CSF samples in the cases. In the frontal cortex, there were also higher levels of N-acetylputrescine (Cohen’s d = 0.84, p < 0.0001) and 1-methylhistamine (d = 0.66, p < 0.0001), and lower levels of aspartic acid (d = -0.53, p = 0.0002) and 3-methylhistidine (d = -0.58, p = 0.0004) in the cases compared to controls, which were statistically significant after adjusting the α level for multiple testing. In the CSF samples, only the cotinine difference between the study groups was significant at the multiple testing-corrected α level (0.0004). Furthermore, there were several other metabolites (e.g., amino acids, xanthines and B-vitamins), which were lower in abundance in the cases as compared to the controls in the frontal cortex (Figure 2) and CSF samples (Figure 3), with p-values between 0.05 and 0.0004. These findings should be considered trends.

**Figure 1:**
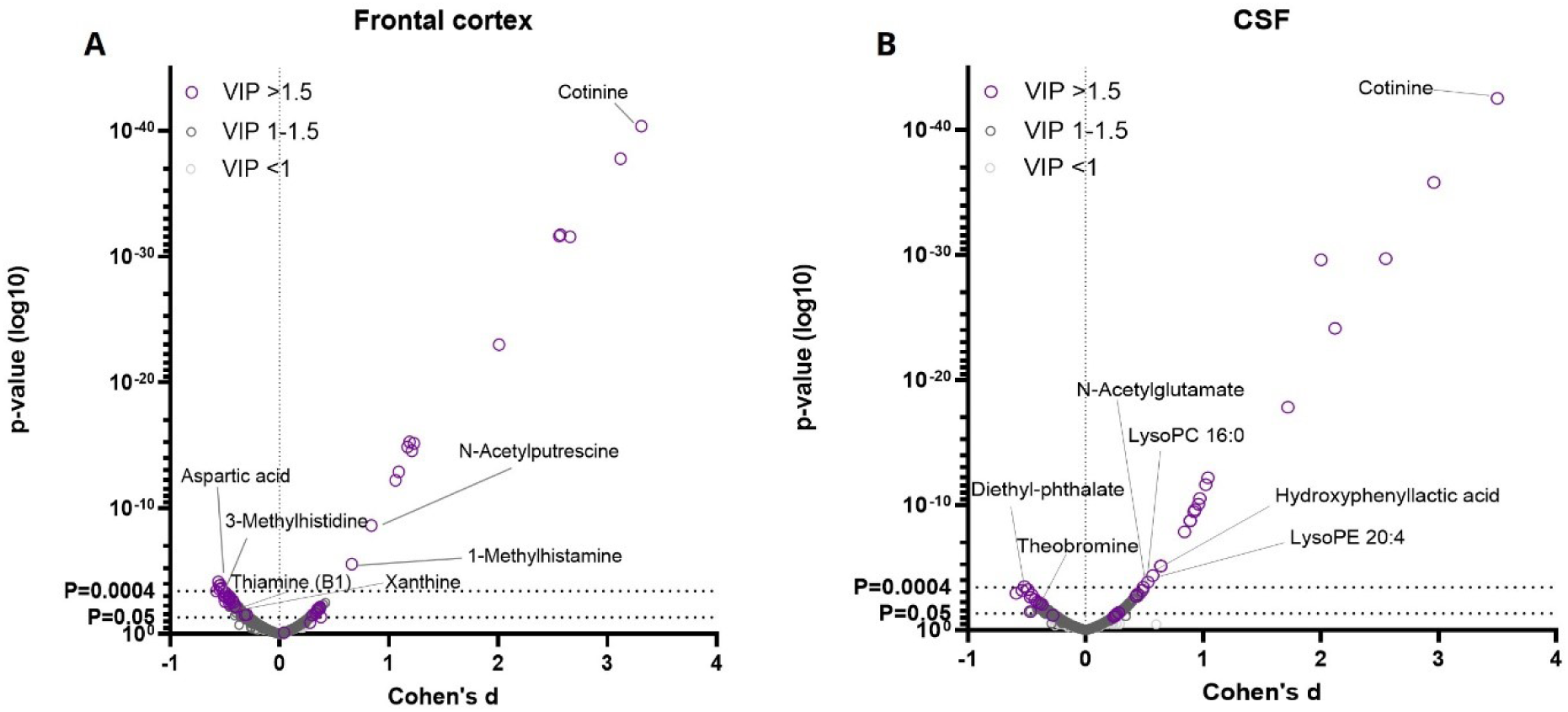
The volcano plots show differences in the frontal cortex and CSF metabolite profile between cases and controls. P-values (Welch’s t-test), Cohen’s d effect sizes and variable importance for projection (VIP, from multivariate PLS-DA analysis) values of all seen molecular features in post-mortem frontal cortex samples are shown in Fig. 1A, and all molecular features in the cerebrospinal fluid samples are shown in Fig. 1B. Some highlighted features were significantly altered between samples of the study groups (multiple testing adjusted α level = 0.0004, Bonferroni’s correction).

**Figure 2:**
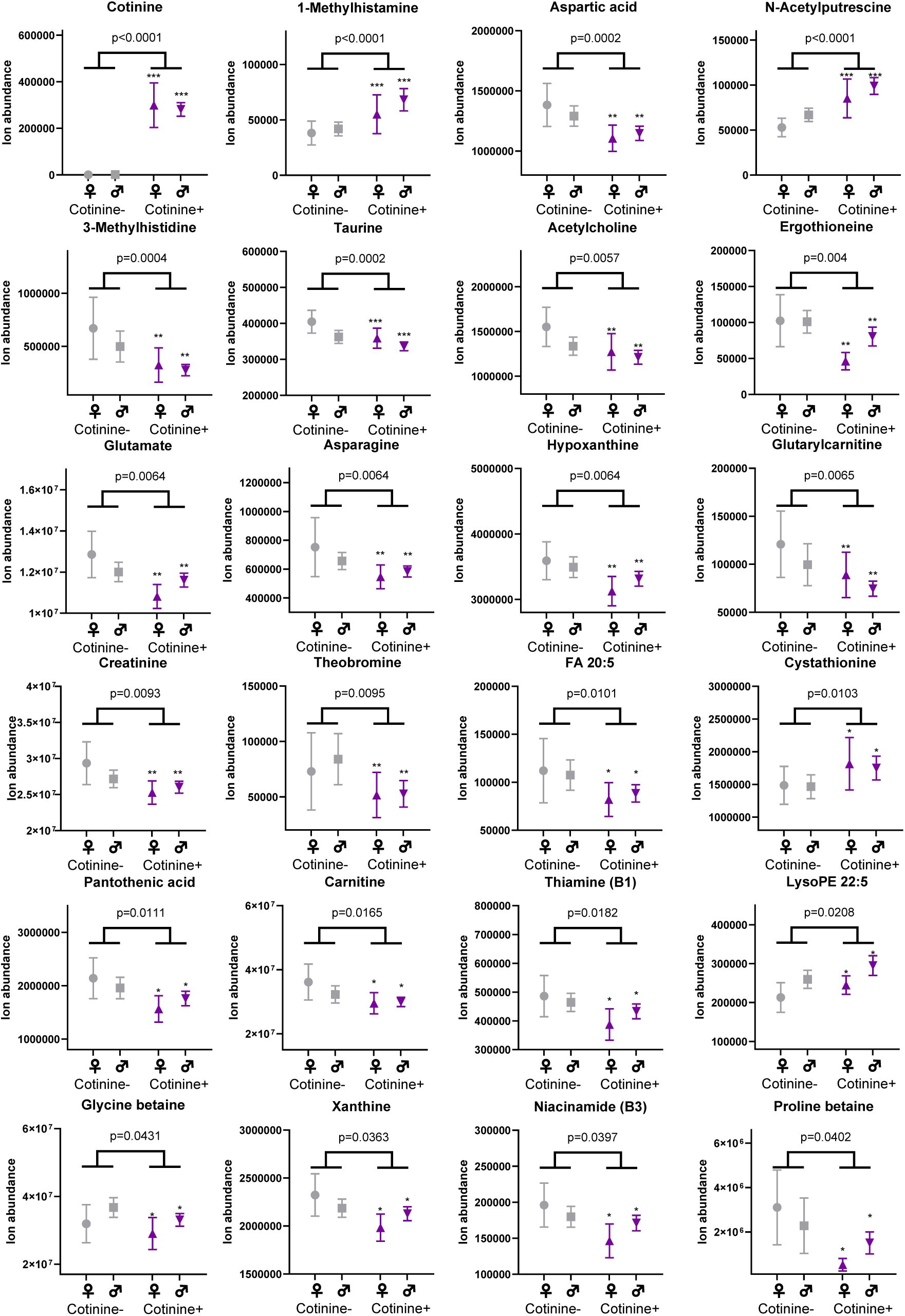
Differences between the study groups in the frontal cortex metabolome. Individual values and group means ± 95% confidence intervals are shown. P-values from Welch’s t-test comparison between all cases and controls is shown as a number. P-values below 0.0004 are significant after correction for multiple testing and p-values between 0.05 and 0.0004 should be considered trends. Welch’s t-test comparisons between same sex cases and controls (male cases vs. male controls, female cases vs. female controls) are shown with asterisks: *, p<0.05; **, p<0.01; ***, p<0.001. (Gray: control vs purple: case)

**Figure 3:**
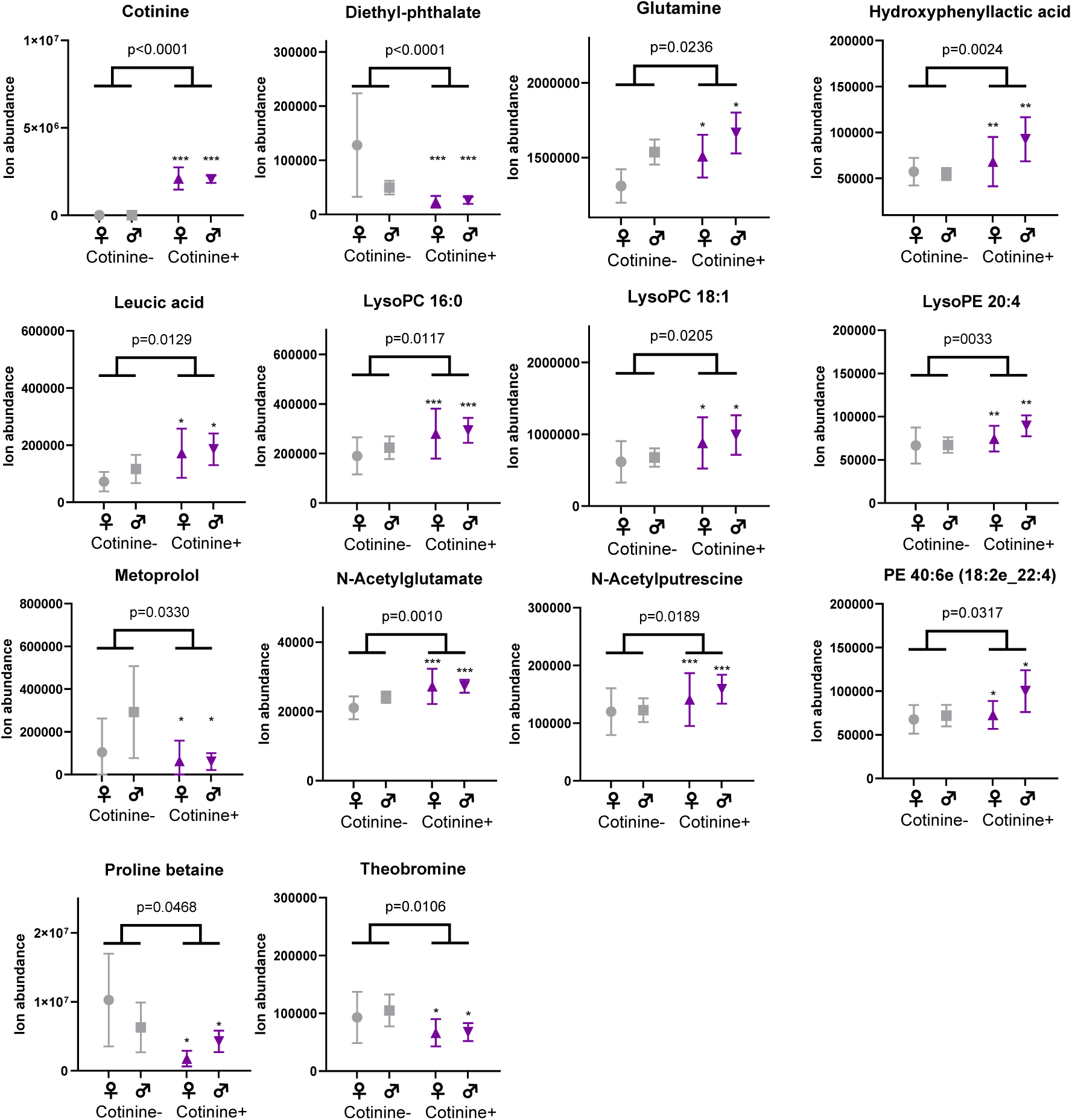
High cotinine levels are associated with alterations in the cerebrospinal fluid metabolome. Individual values and group means ± 95% confidence intervals are shown. P-values from Welch’s t-test below 0.0004 are significant after correction for multiple testing and p-values between 0.05 and 0.0004 should be considered trends. Welch’s t-test comparisons between same sex cases and controls (male cases vs. male controls, female cases vs. female controls) are shown with asterisks: *, p<0.05; **, p<0.01; ***, p<0.001. (Gray: control vs purple: case)

To estimate possible differences between sexes, we performed the statistical analysis separately for both sexes. For all key findings, the direction of the difference in metabolite levels between cases and controls remained the same (Supplementary Table S1). For most of the key findings, the Cohen’s d effect size also remained at relatively similar levels, although some metabolites did show a more pronounced difference between female cases and controls (e.g., lower levels of ergothioneine, d = -1.06, and glutamate, d = -1.08, in the frontal cortex, and higher levels of leucic acid, d = 0.72, and glutamine, d = 0.69, in the CSF, Supplementary Table S1). However, due to reduced statistical power, most p-values were > 0.05 in the comparison between the study groups when analyzing data only from female subjects.

Correlation analysis showed that in the frontal cortex, PMI was positively associated with aspartic acid (r = 0.225, p < 0.001), asparagine (r = 0.389, p < 0.001) and xanthine (r = 0.299, p < 0.001). In the CSF, PMI was positively associated with N-acetylglutamate (r = 0.256, p < 0.001) and negatively associated with LysoPE 22:5 (r = -0.276, p < 0.001, Supplementary Fig 1).

## Discussion

The results of this study show that there are changes in the metabolite profiles of post-mortem human frontal cortex and CSF that are associated with exposure to tobacco or nicotine products, as assessed by high levels of the nicotine metabolite cotinine in the samples. Furthermore, most of the observed differences were similar in both sexes, with a few exceptions, such as low ergothioneine levels observed especially in the female cases.

As expected by study design, the tobacco or nicotine exposure-related metabolites were significantly higher in the cases when compared to the controls. Previous studies have shown that cotinine, a primary metabolite of nicotine, influences the neurotransmitters in the frontal cortex and cerebrospinal fluid^23^. For example, cotinine is effective in the activation of the left prefrontal cortex in people who have quit smoking by enhancing the release of neurotransmitters like dopamine and serotonin, while smokers have reduced activation, which has been linked to differing effects on cognitive processing due to nicotine dependence^24^. However, levels of most neurotransmitters degrade with PMI, which can interfere with the interpretation of results from human post-mortem samples^25^. Glutamate and acetylcholine are generally more stable than serotonin and dopamine, but they can still undergo changes depending on the PMI and storage conditions^25^.

Furthermore, we observed low levels of taurine in the frontal cortex of the cases when compared to the controls, in line with a previous report of decreased levels of taurine in the brain samples of nicotine-exposed mice^26^. In the brain, taurine plays a role in antioxidative function, similar to the microbiota-associated metabolite ergothioneine, which also showed a trend of low levels in the cotinine-positive group in the present study^27,28^. In line with our results, low levels of ergothioneine have also been reported in pregnant women who smoke^17^. These results suggest increased oxidative stress and reduced antioxidative capacity in smoking- or nicotine-exposed brains.

Moreover, the cases had high levels of N-acetylputrescine in the frontal cortex samples when compared to the controls. High N-acetylputrescine levels have been linked to, for example, Parkinson’s disease, glioblastoma and neurodevelopmental problems^29–32^. High N-acetylputrescine levels could also be a marker of increased protein breakdown^33^.

However, this does not seem to be the case here, since another proteolysis marker, 3-methylhistidine, was significantly lower in the cases, and the average PMI was also lower in the cases compared to controls. Therefore, more prolonged bodily degradation does not explain the group difference of N-acetylputrescine.

Additionally, we observed high 1-methylhistamine (a brain-derived metabolite of histamine) levels in the frontal cortex samples from the cases. Previously, high 1-methylhistamine levels have been reported in young adults with a history of high alcohol use, where high 1-methylhistamine levels were associated with low brain gray matter volume^34^. High 1-methylhistamine levels have been proposed to be associated with elevated histamine levels and inflammatory processes in the brain^35^.

Furthermore, the cases had low levels of aspartic acid in the frontal cortex compared to the controls. In the brain, aspartic acid binds to NMDA receptors and is associated with neuroplasticity^36^. There were trends towards lower levels of other neurotransmitters, namely glutamate and acetylcholine, in the frontal cortex samples of the cases. Overall, this suggests that the cases have altered neurochemistry compared to controls. This is consistent with repeated chronic exposure to nicotine, which increases levels of nAChRs and likely leads to a reduction in acetylcholine levels in the brain^7,37,38^. Low levels of these neurotransmitters in the frontal cortex could indicate altered neuroplasticity and activity in the cases. Additionally, a previous study has reported that with nicotine exposure, acetylcholine levels were elevated in the striatum, and glutamate levels were reduced in the hippocampus and striatum, but increased in the nucleus accumbens and prefrontal cortex in mice^26^. In humans, increased circulating levels of glutamate have been reported in smokers^14,16^. However, here we did not see an increase in CSF glutamate levels, and the glutamate and acetylcholine levels were decreased in the frontal cortex. These results indicate that the frontal cortex metabolome is, to some degree, independent of the levels of these metabolites in the blood.

Moreover, we observed trends of decreased levels of several B-vitamins in cases compared to controls in the frontal cortex. Previously, low blood pantothenic acid (B5) is linked to smoking ^13^. These alterations could be due to diet, a notion which is supported by the trends of decreased levels of other metabolites associated with diet like proline betaine and theobromine, seen in the cases.

Limitations of the study include factors like smoking status not being available for all subjects. Additionally, some high cotinine levels could be due to nicotine from other sources, such as snuff or nicotine replacement therapy. Electronic cigarettes, nicotine pouches, or similar products containing high levels of nicotine without tobacco for recreational use were not common in Finland during the period of sample collection (2010– 2015). Therefore, it is presumed that most of the nicotine exposure leading to high cotinine levels in this cohort is due to exposure to tobacco. Furthermore, these results are from a single cohort with a relatively small number of subjects and therefore they should be validated in a larger study with subjects from more diverse backgrounds. Even though there were no significant differences between cases and controls in PMI, some of the results could be influenced by PMI. This is seen in the PMI correlation in the frontal cortex with aspartic acid, asparagine, xanthine, and in the CSF with N-acetylglutamate and LysoPE 22:5. These post-mortem effects increase variation between samples, necessitating larger sample sizes to observe exposure-related effects with adequate statistical power. Despite these limitations, post-mortem samples provide a unique opportunity to obtain high-quality metabolomics data from the human brain^19^. Future studies should investigate how the brain metabolome is altered in subjects exposed to nicotine-only-containing products.

In conclusion, the current study successfully investigated changes in human post-mortem frontal cortex and CSF metabolite profiles associated with tobacco smoking or nicotine exposure. Cases had higher levels of metabolites associated with increased oxidative stress and inflammation in the frontal cortex, and lower levels of neurotransmitters in the frontal cortex. Further studies are needed in several different populations, to broaden the reach of these results.

## Ethics approval statement

The Tampere Sudden Death Study (TSDS) protocol was approved by the Ethics Committee of Pirkanmaa Hospital District (Permission number R09097) and the National Supervisory Authority for Welfare and Health Valvira (Dnro 564/05.01.00.06/2010). This was in accordance with the Declaration of Helsinki.

## Data availability

The data that support the findings of this study are available on request from the corresponding author, OK. The study plan approved by the ethical committee and the participant consent terms preclude public sharing of these sensitive data, even in anonymized form.

## Acknowledgements

We thank Miia Reponen for the excellent technical assistance with the mass spectrometry analyses. The authors want to thank Biocenter Finland and Biocenter Kuopio for supporting their core LC-MS laboratory facility.

